# Unveil the mechanism of Jinzhen Oral Liquid combined with Azithromycin in the treatment of Mycoplasma pneumoniae pneumonia based on Network pharmacology and clinical trials

**DOI:** 10.1101/2024.06.27.24309347

**Authors:** Chengliang Zhong, Shengxuan Guo, Qingyuan Liu, Deyang Sun, Boyang Wang, Siyuan Hu, Xinmin Li, Ying Ding, Bin Yuan, Jing Liu, Long Xiang, Nan Li, Zheng Xue, Yan Li, Yiqun Teng, Rongsong Yi, Shao Li, Rong Ma

**Author notes:** These authors contributed equally: Chengliang Zhong, Shengxuan Guo, Qingyuan Liu, Deyang Sun, Boyang Wang. Corresponding author: E-mail addresses (Rong Ma); (Shao Li).

## Abstract

Mycoplasma pneumoniae pneumonia (MPP) is a common type of pneumonia among school-aged children and adolescents. Jinzhen Oral Liquid (JZOL) and Azithromycin (AZ) are commonly used treatment options in traditional Chinese medicine (TCM) and Western medicine, respectively. There are several clinical and basic research reports on their solo effect against MPP, enabling their combined treatment to become possible. However, the mechanisms and specific pharmacodynamics of their combined therapy remain unclear. In this study, we conducted a mechanistic analysis of the combination of JZOL and AZ based on network target, elucidating their modular network regulatory mechanisms. The modular mechanisms involve four modules, including hormone response, cell differentiation and migration, signal transduction, oxygen and hypoxia response, centered by TNF signaling pathway-mediated regulation. Under the instruction of computational analysis, we conducted a randomized, double-blind, three-armed, parallel-controlled, multicenter clinical study of different doses of JZOL combined with AZ for the treatment of MPP in children. At the study endpoint, the median time to clinical recovery showed statistically significant differences, which were also observed between groups for time to complete fever remission, time to relief of cough/phlegm, effective rate of chest X-ray improvement, and rate of healing of TCM symptoms. During the treatment period, there were no statistically significant differences in the rates of adverse events, serious adverse events, or adverse reactions between the groups. Different doses of JZOL combined with AZ in the treatment of MPP in children have shown the effects of shortening the course of the disease, relieving the symptoms, and improving the prognosis. The research program composed of computational prediction and clinical trials can significantly accelerate the research and development process and identify more effective treatment with good safety, which is worthy of clinical promotion.

## 1. Introduction

Mycoplasma pneumoniae pneumonia (MPP) is a common type of pneumonia in school-aged children and adolescents, accounting for 10-40% of community-acquired pneumonia (CAP) in children [1–3]. Rational treatment, reduction of severe and critical illnesses, and avoidance of death and sequelae are the core and key to the treatment of MPP [4]. Chinese medicine has certain clinical efficacy in different stages of MPP and can play a key role in promoting symptomatic relief and improving the prognosis. The synergistic treatment of Chinese and Western medicine can better promote the recovery of children with MPP and reduce the occurrence of sequelae [5].

Traditional Chinese medicine (TCM) has been extensively used in the treatment of respiratory diseases [6]. Jinzhen Oral Liquid (JZOL) is a proprietary TCM that consists of 8 kinds of Chinese herbs. Pharmacological studies have shown that JZOL can inhibit the activity of Mycoplasma pneumoniae (MP), regulate inflammatory factors induced by different pathogens, reduce pro-inflammatory factors, increase anti-inflammatory factors, regulate anti-inflammatory-promotional imbalance, attenuate inflammatory injury of lungs after infection, attenuate the pathological injury of lungs caused by infections, regulate CD3+/CD4+/CD8+, improve the immune function of T-cells, and regulate the activity of MDA/SOD, anti-peroxide damage, etc. It has good effects of antipyretic, antitussive, reducing sputum production and promoting sputum discharge [7–13]. Previous clinical studies have shown that this product has better efficacy against MPP in children [14–16].

Azithromycin (AZ) is a semi-synthetic nitrogen-15-membered macrolide antibiotic with unique pharmacokinetic characteristics, presenting a multi-chamber model with good tissue permeability and high tissue concentration. Its concentration in cells and tissues can exceed blood concentration by 10-100 times, and the concentration in inflammatory sites is 6 times higher than that in non-inflammatory sites. This optimized internal distribution is highly beneficial for the inhibition and clearance of pathogenic bacteria, and it can also play a unique role in inhibiting protein synthesis in MP. In addition, AZ has a long half-life and significant post-antibiotic effects. Due to the fast uptake and slow release of AZ by tissues, its plasma half-life is close to 70 hours [17]. AZ is a recommended drug in the Chinese Diagnosis and Treatment Guidelines for MPP in Children (2023 Edition) [4].

In order to uncover the mechanism of the combination of JZOL and AZ and provide a new efficient treatment for MPP, a framework containing computational prediction and clinical trial verification was performed based on the theory of network target [18, 19]. Network target is a theory that has been successfully applied in the development and exploration of TCM and modern medicine [20, 21]. Through network-based algorithms and public transcriptomics data, we focused on certain pathways and biological processes, which might depict the key mechanism of the combination of JZOL and AZ. To further evaluate the synergistic effect of JZOL in treating MPP in children to shorten the course of the disease, alleviate the symptoms and improve the prognosis, and to clarify its clinically advantageous dosage, as well as to observe the safety of synergistic application. This study carried out a randomized double-blind, three-armed parallel-controlled, multi-central clinical study to evaluate the differences in time to clinical recovery, time to complete remission of fever, time to remission of cough/sputum, rate of conversion to severe CAP or refractory MPP, chest X-ray efficiency, efficacy of Chinese medicine signs and symptoms, and safety of clinical application with the combined application of AZ and JZOL after a course of 14 days, to provide new ideas for the treatment of MPP.

## 2. Methods and Materials

### 2.1 Clinical trial

#### 2.1.1 Design Overview

This study utilized a stratified cluster-randomized, double-blind, three-arm parallel control, multicenter clinical trial approach. The cases were drawn from children with MPP (pneumonia with wheezing and sputum-heat obstructing lung syndrome or toxic-heat obstructing lung syndrome) treated between November 2018 and December 2020 at twelve clinical research centers, including the First Affiliated Hospital of Tianjin University of Traditional Chinese Medicine, the First Affiliated Hospital of Henan University of Traditional Chinese Medicine, Jiangsu Provincial Hospital of Traditional Chinese Medicine, Beijing Children’s Hospital affiliated with Capital Medical University, Chengdu First People’s Hospital, Shijiazhuang Maternal and Child Health Hospital, Shanghai Hospital of Traditional Chinese Medicine, Suzhou Hospital of Traditional Chinese Medicine, Jiaxing Second Hospital, the Hospital affiliated with Yanbian University, Suzhou Science and Technology Town Hospital, and Liuzhou Maternal and Child Health Hospital. The medication was double-blinded, with the center as the stratification factor. The patients were cluster-randomized in a 1:1:1 ratio into three groups: double-dose of JZOL group, regular-dose group, and placebo group, using SAS software to generate a random number table. The blinding of the medication and emergency letters were handled by third-party personnel not involved in this study. The research has been registered with the China Clinical Trials Registry (registration number: ChiCTR1800019007) and has been approved by the Medical Ethics Committee of the First Affiliated Hospital of Tianjin University of Traditional Chinese Medicine, which is the ethical review of the leading unit’s medical ethics committee (ethics approval number: TYLL2017[Y]020).

#### 2.1.2 Diagnosis and Pattern Differentiation Criteria

The Western medical diagnosis criteria for children with MPP are based on the “Expert Consensus on the Diagnosis and Treatment of Mycoplasma Pneumoniae Pneumonia in Children” (2015) and “Zhu Futang Practice of Pediatrics” (8th Edition) [1, 22]. The TCM criteria for diagnosing pneumonia with wheezing and sputum-heat obstructing lung syndrome or toxic-heat obstructing lung syndrome are derived from the “Guidelines for the Diagnosis and Treatment of Common Pediatric Diseases in Traditional Chinese Medicine - Pneumonia with Wheezing” (2012) [23] published by the Chinese Association of Traditional Chinese Medicine.

#### 2.1.3 Patient Enrollment

Inclusion Criteria: (1) Children who meet the Western medical diagnostic criteria for MPP and the TCM pattern differentiation standards; (2) Aged 2 to 13 (under 14) years; (3) Etiological diagnosis, using the Particle Agglutination (PA) assay to test for serum IgM and IgG mixed antibodies, with a single MP antibody titer of ≥1:160, or a positive result from a single measurement of MP-IgM by Enzyme-Linked Immunosorbent Assay (ELISA); (4) The informed consent process complies with regulations, and the legal guardian or the child subject (≥8 years old) jointly signs the informed consent document.

Exclusion Criteria: (1) Those meeting the criteria for severe CAP [24]; (2) Those meeting the criteria for refractory MPP [1]; (3) Those with diseases needing differentiation from MPP, such as pulmonary tuberculosis, bacterial or viral pneumonia, hospital-acquired pneumonia, and other pathogenic microbial pneumonias; (4) Those with underlying conditions such as primary immunodeficiency diseases, acquired immune deficiency syndrome, congenital respiratory tract anomalies, abnormal pulmonary development, aspiration pneumonia, or malignant pulmonary tumors; (5) Those with severe malnutrition or rickets, as well as severe primary diseases of the heart, brain, liver, kidney, and hematopoietic systems; (6) Those diagnosed with MP infection in the past three months; (7) Those with an allergic constitution (allergic to two or more substances), or allergies to macrolide antibiotics and any components of JZOL; (8) Those deemed unsuitable for the study by the researchers.

Drop-out Criteria: (1) Those who experience an allergic reaction or serious adverse event and are advised by a doctor to discontinue the trial; (2) Those who continue to have a high fever (≥39°C) 72 hours after first medication, or whose condition worsens at any time post-medication, showing symptoms such as shortness of breath, difficulty breathing, or cyanosis, should be promptly withdrawn from the trial to protect the subject; (3) Subjects with poor compliance (medication compliance <80% or >120%), or who autonomously change medications or use prohibited Western or Chinese medicines during the trial; (4) Those who are unblinded for any reason; (5) Those who severely violate the inclusion or exclusion criteria, who should not have been randomized; (6) Patients who, for any reason, are unwilling or unable to continue with the clinical trial and request to withdraw through their attending physician; (7) Subjects who do not explicitly request to withdraw but are lost to follow-up or no longer accept the trial medication and testing.

#### 2.1.4 Interventions

All three groups were administered AZ as the baseline treatment. Injectable AZ (Hisun, T54639) was administered at a dose of 10 mg/(kg·d), diluted in 250-500 ml of 5% glucose solution, and given by intravenous drip. After 5 days of continuous use, the medication was discontinued for 3 days, then switched to an oral dosage (Hisun, T95018) form for another 3 days. The therapeutic medication used was JZOL (produced by Jiangsu Kangyuan Pharmaceutical Co., Ltd., specification: 10ml/per unit, Chinese medicine approval number Z10970018, 171204), administered as follows: Double-dose group (JZOL), regular-dose group (50% concentration of JZOL), or placebo group (JZOL placebo), taken orally, 2-3 years old, 20 ml per dose, twice a day; 4-7 years old, 20 ml per dose, three times a day; 8-13 years old, 30 ml per dose, three times a day. Actual medication dosages (original specifications) for each dose group: (1) Double-dose group, 2-3 years old, 20 ml per dose, twice a day; 4-7 years old, 20 ml per dose, three times a day; 8-12 years old, 30 ml per dose, three times a day. (2) Regular-dose group, 2-3 years old, 10 ml per dose, twice a day; 4-7 years old, 10 ml per dose, three times a day; 8-13 years old, 15 ml per dose, three times a day. (3) Placebo group, placebo administered according to age group. The treatment period was 14 days. Treatment could be terminated at any time after 7 days or more if clinical recovery was achieved.

Concomitant Medication Rules: During the trial, the use of steroids or other Chinese medicines with similar effects to the trial medication was prohibited. If the axillary temperature reached ≥38.5℃, paracetamol (Tylenol, 180310135) at a dose of 5-10 mg/(kg·dose) could be administered orally. If a bacterial infection was confirmed, other antibiotics could be used in combination. The rational use of antibiotics should refer to the “Basic Principles for the Clinical Application of Antimicrobial Agents (2015)” [25], with adjustments to the antibiotic treatment plan based on early identification of the pathogen and results from drug sensitivity tests. Empirical antibiotic treatment should be initiated before the results of bacterial culture and drug sensitivity tests are available.

#### 2.1.5 Effectiveness evaluation

Observational Indicators and Timing: (1) Clinical recovery time, assessed at the end of the trial, calculated in days; (2) Complete fever resolution time, with body temperature measured and recorded every 24 hours from baseline and post-treatment, evaluated at the end of the trial, calculated in days; (3) Relief time for cough and sputum, with symptom severity recorded every 24 hours from baseline and post-treatment, evaluated at the end of the trial, calculated in days; (4) Rate of progression to severe CAP or refractory MPP, evaluated at the end of the trial; (5) Chest X-ray efficacy, examined at baseline and the end of the trial, evaluated at the end of the trial; (6) Efficacy of TCM syndrome differentiation, with syndrome scores recorded at baseline, day 7 of treatment, and the end of the trial, evaluated at the end of the trial. Clinical recovery time is the primary evaluation metric.

Efficacy Evaluation Criteria and Indicator Definitions: (1) TCM Syndrome Scoring Standards: Primary symptoms such as fever, cough, thick sputum difficult to expectorate, and wheezing are graded as normal, mild, moderate, and severe, with scores of 0, 2, 4, and 6, respectively. Secondary symptoms such as sore throat, dry nostrils, flushed face, restlessness, thirst, poor appetite, constipation, and dark scanty urine are also graded as normal, mild, moderate, and severe, with scores of 0, 1, 2, and 3, respectively. (2) Definition of “Complete Defervescence”: Body temperature (axillary) below 37.3°C, maintained for 24 hours or more. (3) Definition of “Relief of Cough or Sputum”: “Cough or sputum” score reduced to mild or normal, not affecting daily activities, and maintained for 24 hours or more. (4) Definition of “Clinical Cure”: Simultaneous fulfillment of “Complete Defervescence” and “Relief of Cough and Sputum”. (5) Definition of Severe Pneumonia: Refer to the “Management Guidelines for Community-Acquired Pneumonia in Children (2013 Revision)” [24]. Based on a diagnosis of MPP, severe pneumonia is indicated by poor general condition, refusal to eat or signs of dehydration, disturbed consciousness, cyanosis, difficulty breathing (moaning, flaring of the nostrils, use of accessory muscles to breathe), pleural effusion, significantly increased respiratory rate (infants RR > 70/min, older children RR > 50/min), involvement of multiple lung lobes or ≥2/3 lung infiltration, pulse oximetry saturation ≤92%, or serious extrapulmonary complications. Any of these conditions indicate severe pneumonia. (6) Definition of Refractory MPP: Refer to the “Expert Consensus on the Diagnosis and Treatment of Mycoplasma Pneumoniae Pneumonia in Children (2015)” [1]. Refractory MPP is defined as MPP that does not improve or worsen after at least 7 days of standard treatment with macrolide antibiotics, with continued fever and worsening pulmonary imaging. (7) Chest X-Ray Efficacy Evaluation Standards: Normal, complete absorption of bilateral pneumonic changes; Significant Improvement, major absorption of bilateral pneumonic changes; Improvement, partial absorption of bilateral pneumonic changes; No Improvement, no change or worsening of bilateral pneumonic changes. Effective, indicated by Normal + Significant Improvement + Improvement. (8) TCM Syndrome Efficacy Evaluation Standards: Clinical Cure, disappearance or near disappearance of TCM clinical symptoms, syndrome score reduction ≥95%; Marked Effectiveness, significant improvement in TCM clinical symptoms, syndrome score reduction ≥70% and <95%; Effective, improvement in TCM clinical symptoms, syndrome score reduction ≥30% and <70%; Ineffective, no significant improvement in TCM clinical symptoms or worsening, syndrome score reduction <30%.

#### 2.1.6 Safety Evaluation

Clinical adverse events (symptoms, signs, disease syndromes, physicochemical abnormalities requiring intervention)/reaction incidence; Vital signs: such as blood pressure, respiration, body temperature, heart rate, etc.; Laboratory tests, such as routine blood and urine tests, liver and kidney function, electrocardiogram, etc. The incidence of clinical adverse events/reactions is the primary safety evaluation indicator.

#### 2.1.7 Statistical Analysis

Statistical analysis was conducted using SAS software, version 9.4, with all tests being two-sided and α set at 0.05. All confidence intervals (CI) are stated with 95% credibility. For quantitative data, describe the number of cases, mean and standard deviation (x±s) or median and interquartile range [M (Q1, Q3)]; group comparisons are made using ANOVA or Kruskal-Wallis test (H test). For qualitative data, describe the number of cases and percentages; group comparisons are made using the chi-square test or Fisher’s exact test. For survival data, describe the median survival time; comparisons between groups are made using the Log-rank test. Missing efficacy indicators are handled using the last observation carried forward method. Subgroup analysis based on syndrome types is conducted.

### 2.2 Network target analysis

#### 2.2.1 Identification of the compounds in JZOL

1 mL of JZOL was applied to an activated solid-phase extraction column (SPE C18 column). The pure methanol eluate was collected and evaporated to dryness under a nitrogen stream. The residue was reconstituted in 1 mL of 50% methanol, and the solution was centrifuged at 14,000 rpm for 10 minutes. The supernatant obtained served as the test solution. The test solution was filtered through a 0.22 μm microporous membrane, and the filtrate was subjected to mass spectrometry analysis. Following sample preparation, data acquisition was performed using UPLC-Q/TOF-MS. The acquired data were imported into the MassLynx V4.1 data processing software for analysis. A total of 90 compounds were identified in JZOL, including 39 flavonoids, 13 alkaloids, 12 triterpene saponins, 9 bile acids, 7 diterpenes, 6 anthraquinones, and 4 other types of compounds.

#### 2.2.2 Target prediction and validation for AZ and compounds in JZOL

The targets of AZ and compounds in JZOL were predicted using the DrugCIPHER algorithm [26]. DrugCIPHER is a network-based computational framework that integrates pharmacological and genomic spaces to predict drug targets on a genome-wide scale. Further, we calculated the holistic targets of JZOL based on our previous statistical strategy [27]. To confirm the accuracy of the result, literature mining was conducted to validate the predicted targets. For each compound in JZOL, we searched PubMed for reported bio-entities such as genes, RNA, or proteins by looking for their names in abstracts and counted the number of relevant papers. We then downloaded these abstracts and extracted the mentioned bio-entities. The predicted targets for each compound were matched with the mined bio-entities.

#### 2.2.3 Differential gene analysis and key pathway identification of MPP

Differential gene analysis of MPP was conducted using data from the GSE179051 [28] dataset, which includes normal HeLa cells and HeLa cells infected with MP. The DESeq2 package was utilized to analyze the gene expression count matrix, with the thresholds set at p-adjust value < 0.05 and |log2FoldChange| > 1. This analysis identified genes that are significantly upregulated or downregulated in MP-infected cells compared to normal cells. Subsequently, enrichment analysis was performed using the GO and KEGG databases to identify significantly enriched pathways, which were then designated as key pathways of MPP.

#### 2.2.4 Network target analysis of JZOL combined with AZ in the treatment of MPP

To elucidate the intervention mechanism of JZOL combined with AZ in the treatment of MPP, the total targets were enriched in MPP-related pathways to identify the key pathways involved in the intervention of MPP. Additionally, an overlap analysis of the total targets of JZOL and AZ was performed based on the biological functional modules constituted by the pathways. Based on this foundation, a database search was conducted to identify the key biomolecular mechanisms underlying the combined treatment of JZOL and AZ for MPP.

#### 2.2.5 Validation on the perturbation set of JZOL compounds

Based on the perturbation set of TCM compounds obtained from ITCM [29], the perturbation genes of key compounds contained in JZOL were identified. First, compounds present in JZOL were extracted from the database. Differential gene expression analysis was then performed between the perturbed transcriptome and the normal transcriptome of these compounds to identify significantly perturbed genes.

## 3. Results

### 3.1 Network target analysis revealing the regulatory role of JZOL combined with AZ in the treatment of MPP

#### 3.1.1 Biological effective profile prediction

Based on UPLC-Q/TOF-MS, 90 compounds contained in JZOL were identified. Using the DrugCIPHER algorithm, the target profile of each compound in JZOL, as well as AZ, was calculated. Additionally, statistical methods were employed to determine the total target profile of each herb in JZOL and the total target profile of JZOL combined with AZ. It was observed that there was a significant overlap in the targets of each herb in JZOL, particularly in Scutellaria baicalensis (Huangqin), Rheum palmatum (Dahuang), and Glycyrrhiza uralensis (Gancao), demonstrating the synergistic effects of the herbs in JZOL (Figure 1B). The accuracy of the predicted compound targets was validated through literature mining. Statistical analysis revealed that the key quality control compounds of JZOL and their corresponding predicted targets co-appeared in the literature with a probability of 80%-95% (Figure 1A). This indicates that the predicted compound targets are closely related to the molecular mechanisms discussed in the literature, confirming their reliability.

**Figure 1.**
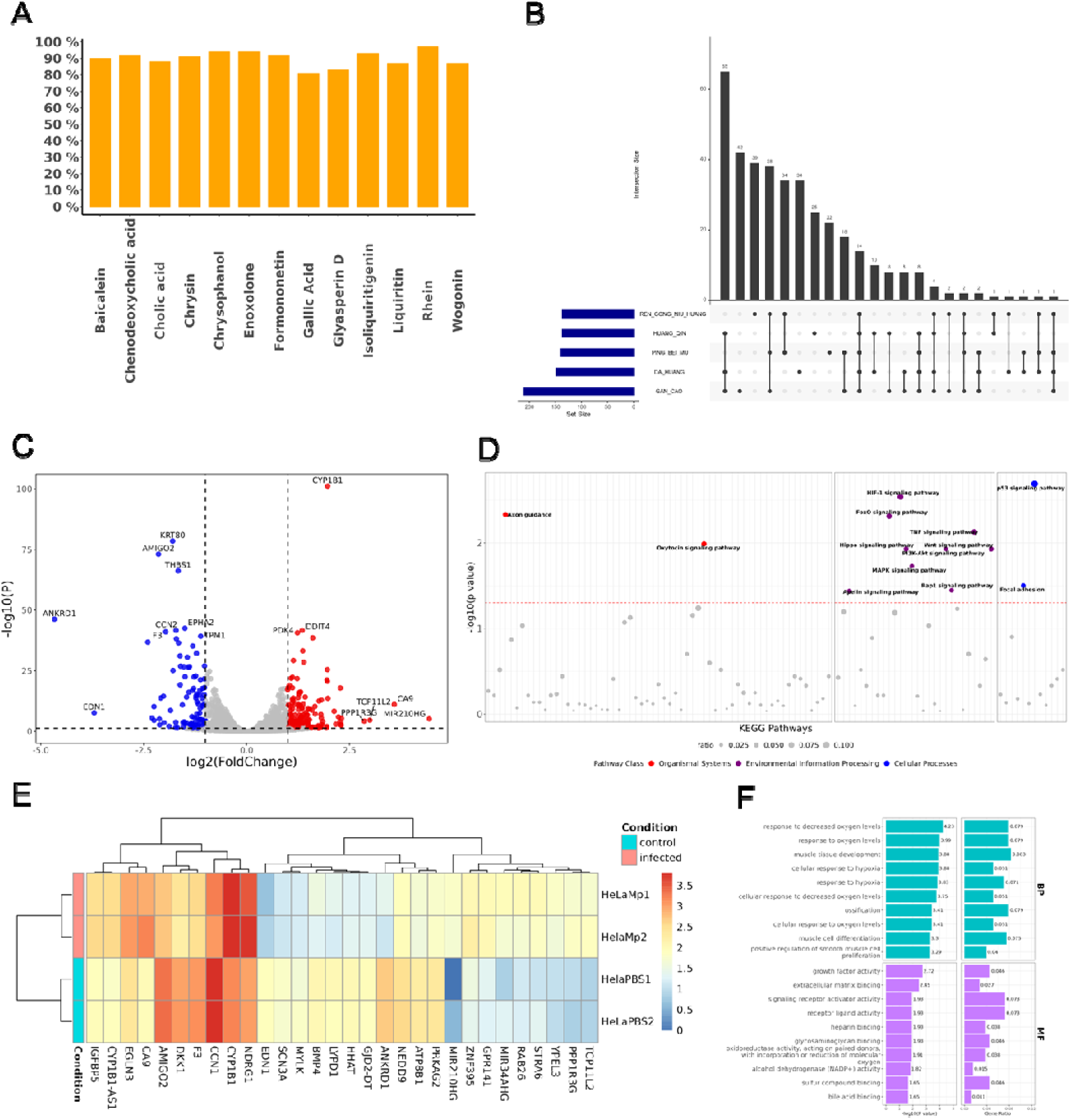
Biological effective profile of JZOL and mechanism of MPP. (A) Literature validation of predicted targets of JZOL quality control compounds. (B) Target overlap of each herb in JZOL. (C) Differential gene analysis of MP-infected cell lines. (D) KEGG pathway enrichment of MP-infected cell lines. (E) Clustering of differential genes in MP-infected cell lines. (F) GO pathway enrichment of MP-infected cell lines

#### 3.1.2 Transcriptomics revealing mechanism of MPP

Through differential gene analysis of the transcriptomes of MP-infected cell lines and normal cell lines, 287 genes significantly upregulated or downregulated in association with MPP were identified (Figure 1C). The hierarchical clustering of these genes revealed modular characteristics (Figure 1E). Further enrichment analysis of these differential genes on KEGG and GO biological pathways indicated that MPP-related biological functional pathways were concentrated in modules such as organismal systems, environmental information processing, cellular processes, and oxygen response (Figure 1F).

#### 3.1.3 Modular regulation of JZOL combined with AZ against MPP

Our analysis revealed that the mechanism of JZOL combined with AZ against MPP involves a total of four modules, and they are as follows, hormone response, cell differentiation and migration, signal transduction, oxygen and hypoxia response. Some pathways and biological processes associated with these modules had been extensively documented in the literature and played crucial roles in MPP. In the hormone response module section, it is confirmed that corticosteroid could effectively initiate the rapid improvement of clinical symptoms and chest radiographic findings in hospitalized MPP patients [30–32]. In the cell differentiation and migration module, mycoplasma colonization on the apical surface of airway epithelium, and the epithelial furrows as a part of a coordinated epithelial remodeling during persistent MP infection may involve epithelial-to-mesenchymal transition (EMT) [33].

In the signal transduction section, it involves many signaling pathways, such as the TNF signaling pathway, PI3K-AKT signaling pathway, MAPK signaling pathway, and WNT signaling pathway, all of which have been proven to play important roles in MPP. It was found that serum TNF-α levels in children with refractory M. pneumoniae pneumonia (RMPP) were significantly higher than those in children with non-RMPP [34]. The levels of cytokines related to the PI3K-AKT signaling pathway in severe Mycoplasma pneumoniae pneumonia (SMPP) were also found higher than those in the healthy control group. The MAPK signaling pathway, as a signaling pathway that produces excessive pro-inflammatory cytokines, can be released by lipid-associated membrane proteins (LAMPs) binding to Toll-like receptor 2 (TLR2) on immune cells during the occurrence of MPP [35]. The WNT signaling pathway can also be activated after MPP infection, thereby inhibiting the secretion of inflammatory factors to quickly clear pathogens [36]. In the oxygen and hypoxia response section, MPP with hypoxia was more likely to be accompanied by atelectasis, pleural effusion, and aggravated in a short period [37, 38].

#### 3.1.4 Network target of JZOL combined with AZ against MPP

The TNF signaling pathway related cytokines are the first line of defense against MPP secreted by airway epithelial cells, which induce the recruitment of inflammatory cells, leading to local inflammation, airway remodeling, airflow obstruction, emphysema, and a decline in lung function [39]. Children with refractory MPP often have higher serum levels of TNF-α [34, 40], and TNF-α was highlighted to be prominent to evaluate the MPP [41]. Besides, the expression levels of TNF signaling pathway related factors could be reduced by the treatment of AZ during MPP [42, 43].

Firstly, based on the public transcriptomics dataset ITCM, 8 compounds from 5 different herbs in JZOL were selected. By analyzing the gene expression profiles after the perturbation by these compounds, eight significantly affected genes were identified (Figure 2 C). The results indicate that the compounds from each herb notably intervene with these genes. Certain biomolecules in these pathways have been reported to play important roles in MPP. MP can bind to TLR2 and activate MAPKs, ERK1/2, JNK, etc., inducing significant time-dependent nuclear translocation of NF-κB and phosphorylation of PI3K/Akt in airway epithelial cells, leading to a significant increase in IL-8 production [44, 45]. The activation of NF-κB may be PI3K/Akt independent or dependent; the latter has received support from many studies [46, 47]. In addition, the increase of TNF-α in the submucosa during MP infection interacts with TNF-R on mast cell, leading to the release of a large amount of pre-existing and newly synthesized mediators. Many mediators from degranulated mast cells, including cytokines, chemokines, lipid mediators, and histamine, may play key roles in the pathophysiology of MPP. It is precisely one or several synergistic media that cause increased mucus production and airway hyperresponsiveness [48, 49].

**Figure 2.**
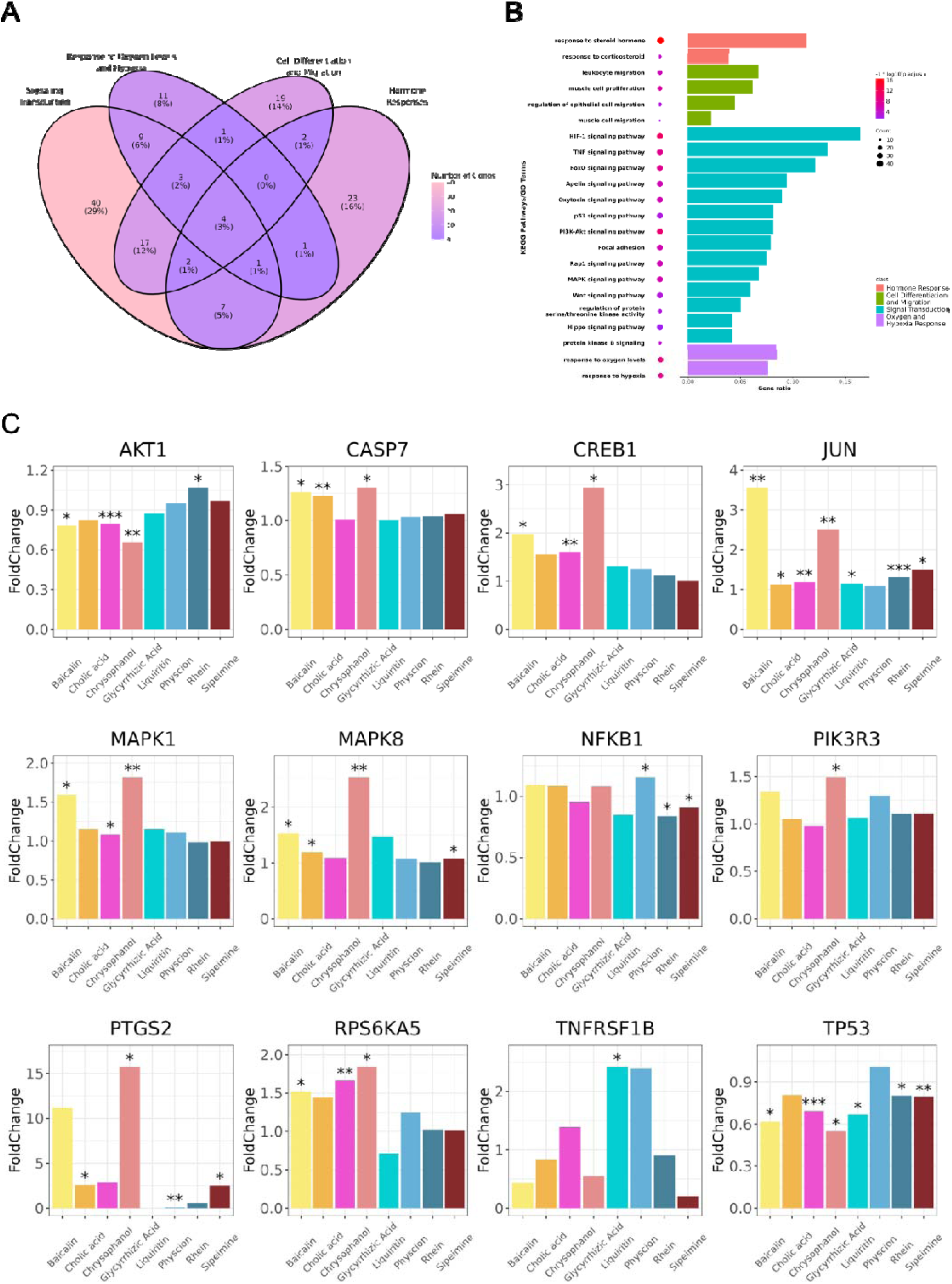
Modular regulation of JZOL combined with AZ against MPP. (A) Overlap analysis of the total targets of JZOL and AZ based on the biological functional modules. (B) Biological functional enrichment of the total targets of JZOL and AZ in key MPP pathways. (C) Gene perturbation results by JZOL compounds on public transcriptome dataset.

As a significantly enriched pathway by the targets of the combination of JZOL and AZ, the TNF signaling pathway has crosstalk with other pathways potentially regulated by the combination, like PI3K-Akt and MAPK signaling pathways. We further focused on these pathways and their crosstalk, predicting the key regulatory mechanism of the combination of JZOL and AZ. With the combination of [50] target analysis and public transcriptomics data validation, we depicted the key mechanism of the combination of JZOL and AZ in the treatment of MPP (Figure 3C). Specifically, this combination impacts the TNF signaling pathway, involving TNFR1 and TNFR2, leading to the activation of various kinases such as NF-κB, ERK1/2, and JNK1/2. One effect of this activation is the regulation of apoptosis through CASP7. Simultaneously, through the MAPK signaling pathway, biomolecules such as AP-1 and CREB are activated, which promote processes like leukocyte recruitment, extracellular matrix remodeling, and the synthesis of inflammatory mediators. These processes collectively enhance cell survival and reduce inflammation. Furthermore, the activation of the PI3K-Akt pathway influences the expression of proteins like p53, thereby further enhancing cell survival. Overall, the combined action of JZOL and AZ strengthens the immune response, reduces inflammation, and promotes tissue repair, thereby improving the treatment efficacy for MPP.

**Figure 3.**
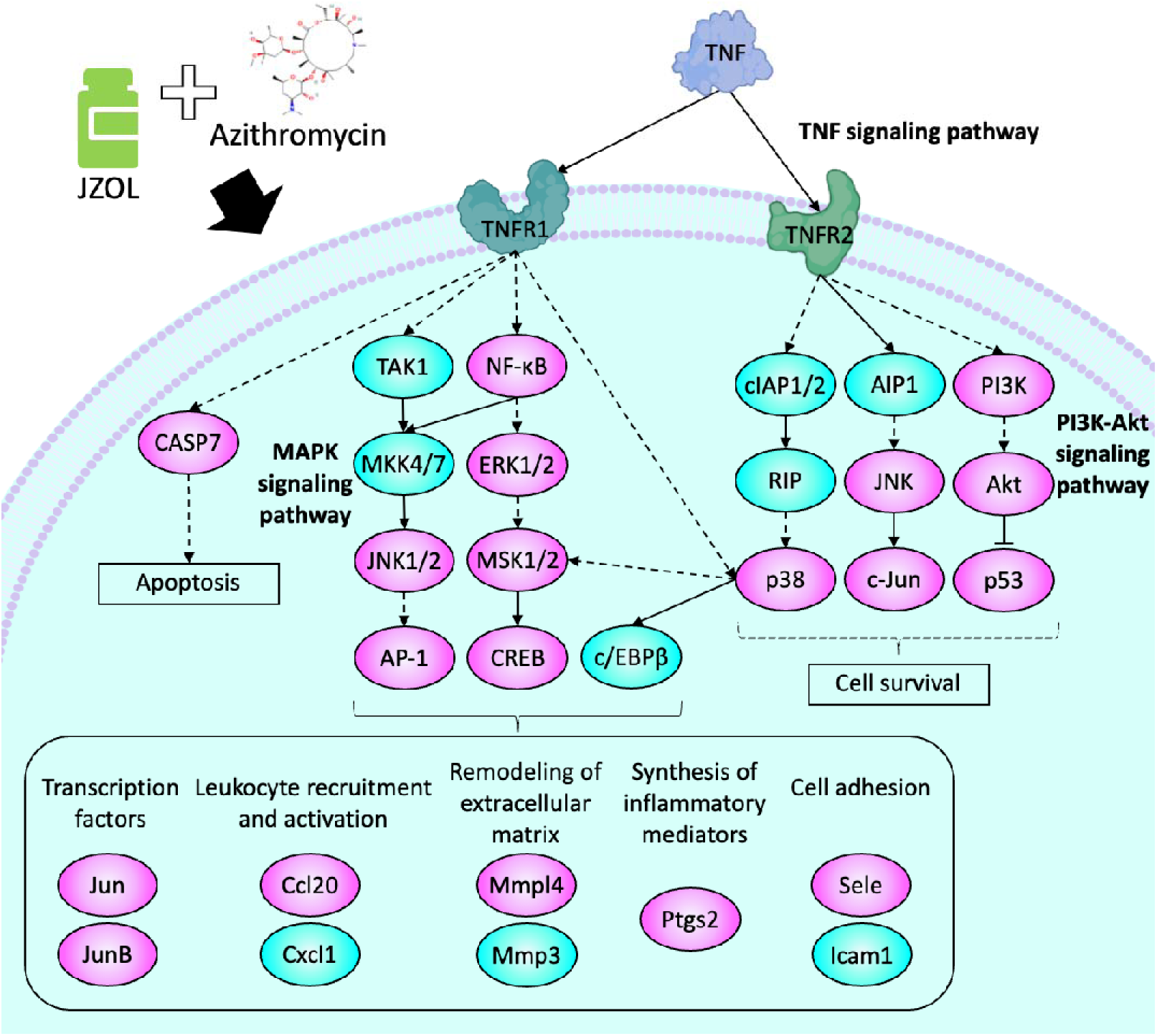
Key mechanism diagram of the combination of JZOL and AZ in the treatment of MPP. By intervening in the TNF signaling pathway, MAPK signaling pathway, and PI3K-Akt signaling pathway, the therapeutic regimen of JZOL combined with AZ impacts cell survival and apoptosis, transcription factors, leukocyte recruitment and activation, extracellular matrix remodeling, inflammatory mediator synthesis, and cell adhesion.

**Figure 4.**
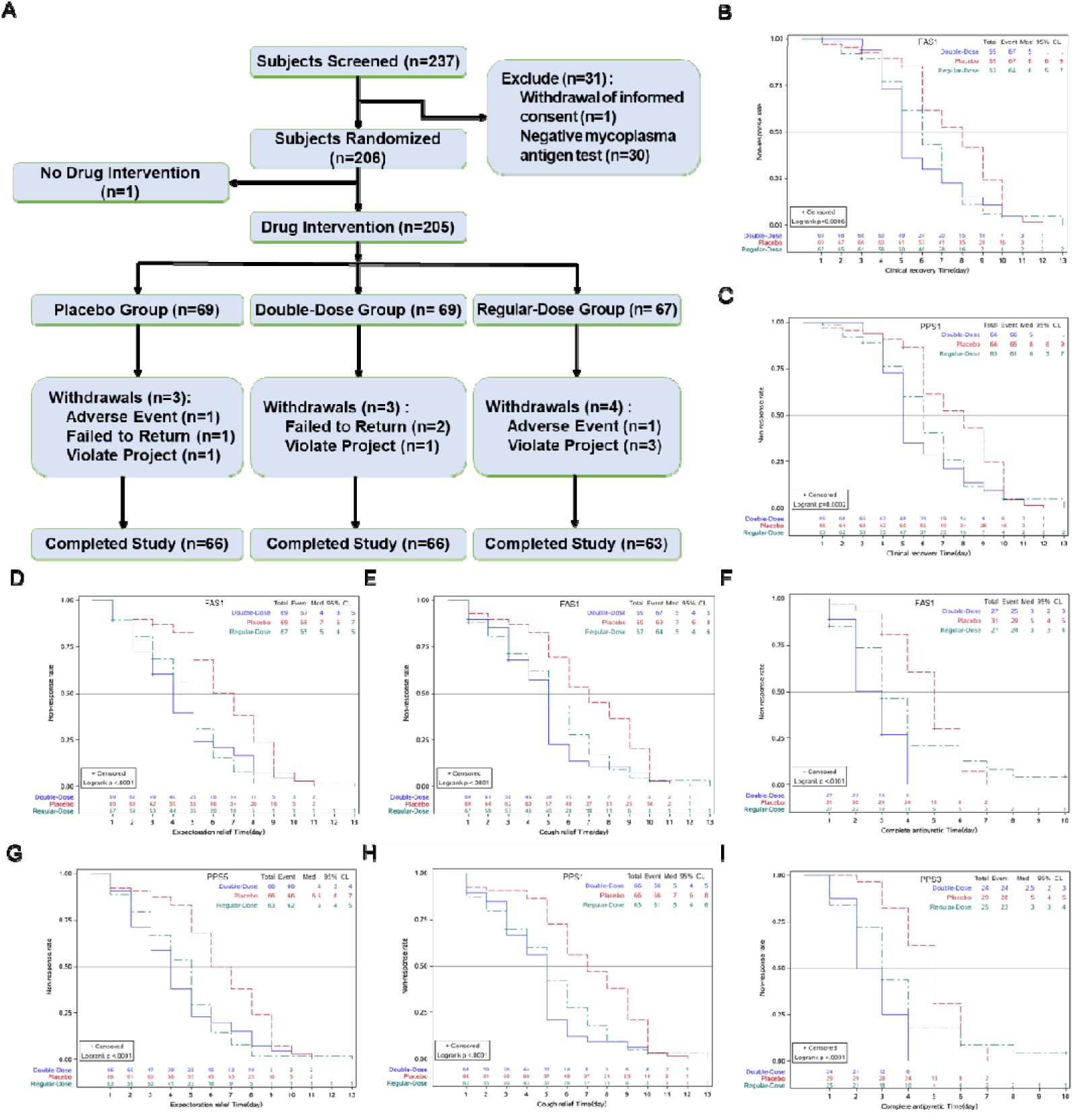
Primary clinical outcomes. (A) Clinical trial patient enrollment and grouping diagram. (B) FAS analysis of clinical recovery time. (C) PPS analysis of clinical recovery time. (D) FAS analysis of expectoration relief time. (E) FAS analysis of cough relief time. (F) FAS analysis of complete antipyretic time. (G) PPS analysis of expectoration relief time. (H) PPS analysis of cough relief time. (I) PPS analysis of complete antipyretic time.

#### 3.1.5 Participant Characteristics

A total of 206 subjects were enrolled in this trial, including 69 in the double-dose group, 68 in the regular-dose group, and 69 in the placebo group. One subject did not take the trial medication and was not included in the analysis set. A total of 205 subjects were included in the Full Analysis Set (FAS), 195 in the Per Protocol Set (PPS), and 205 in the Safety Set (SS). Reasons for exclusion from the PPS included adverse events, voluntary withdrawal, and use of prohibited medications during the treatment course.

All children included in the FAS analysis had baseline demographic data (gender, ethnicity, age, height, weight) and disease-related conditions (duration of fever, highest body temperature in the 24 hours before diagnosis, immediate body temperature at enrollment, medical history, family history, allergy history, pre-enrollment comorbidities and treatments, TCM syndrome types, concurrent infections) compared between groups. There were no statistically significant differences, ensuring comparability. Efficacy-related indicators, including baseline TCM syndrome scores and individual symptom scores, also showed no statistically significant differences between groups (Table 1).

**Table 1.**
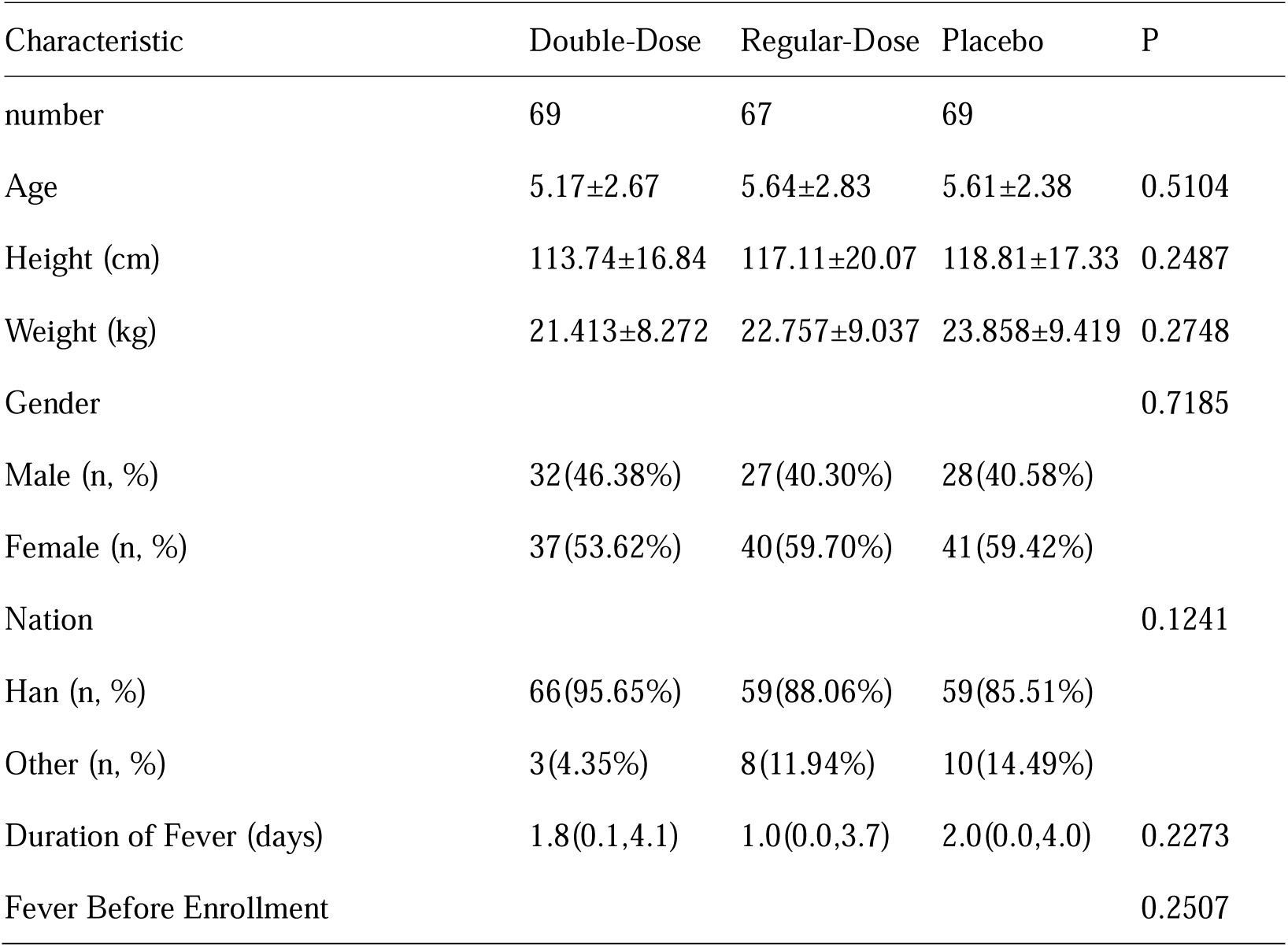
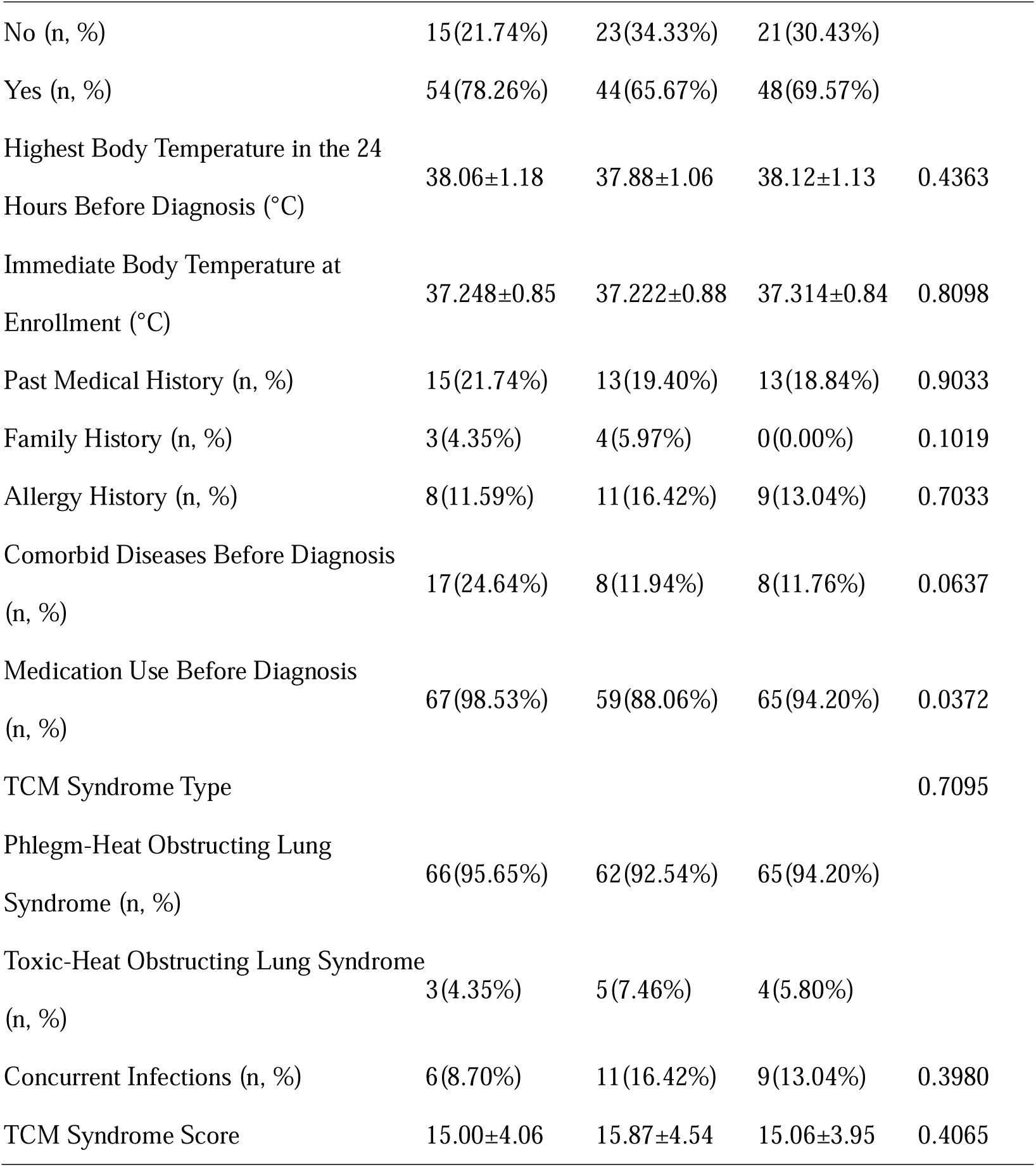
Comparison of three groups of patients at baseline.

**Table 2.**
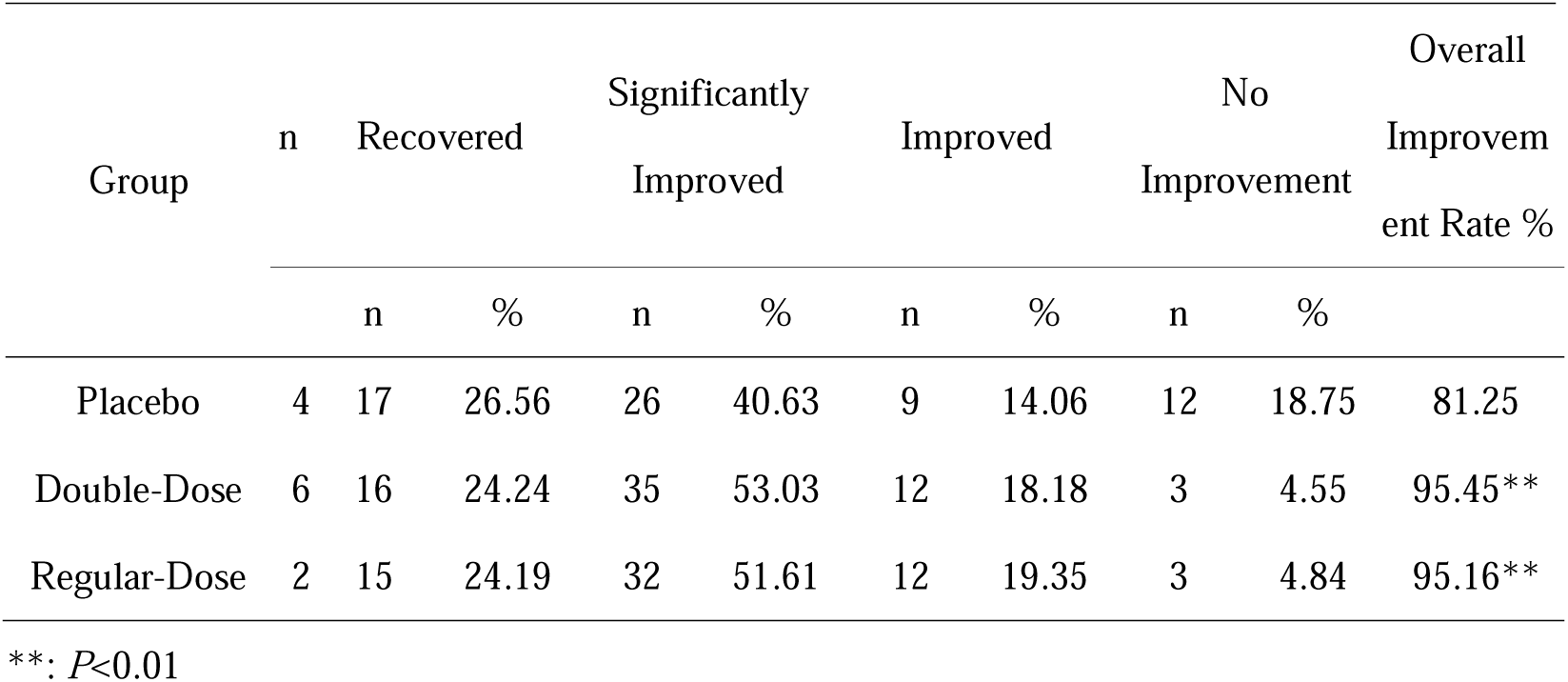
Comparison of chest X-ray improvement in three groups.

**Table 3.**
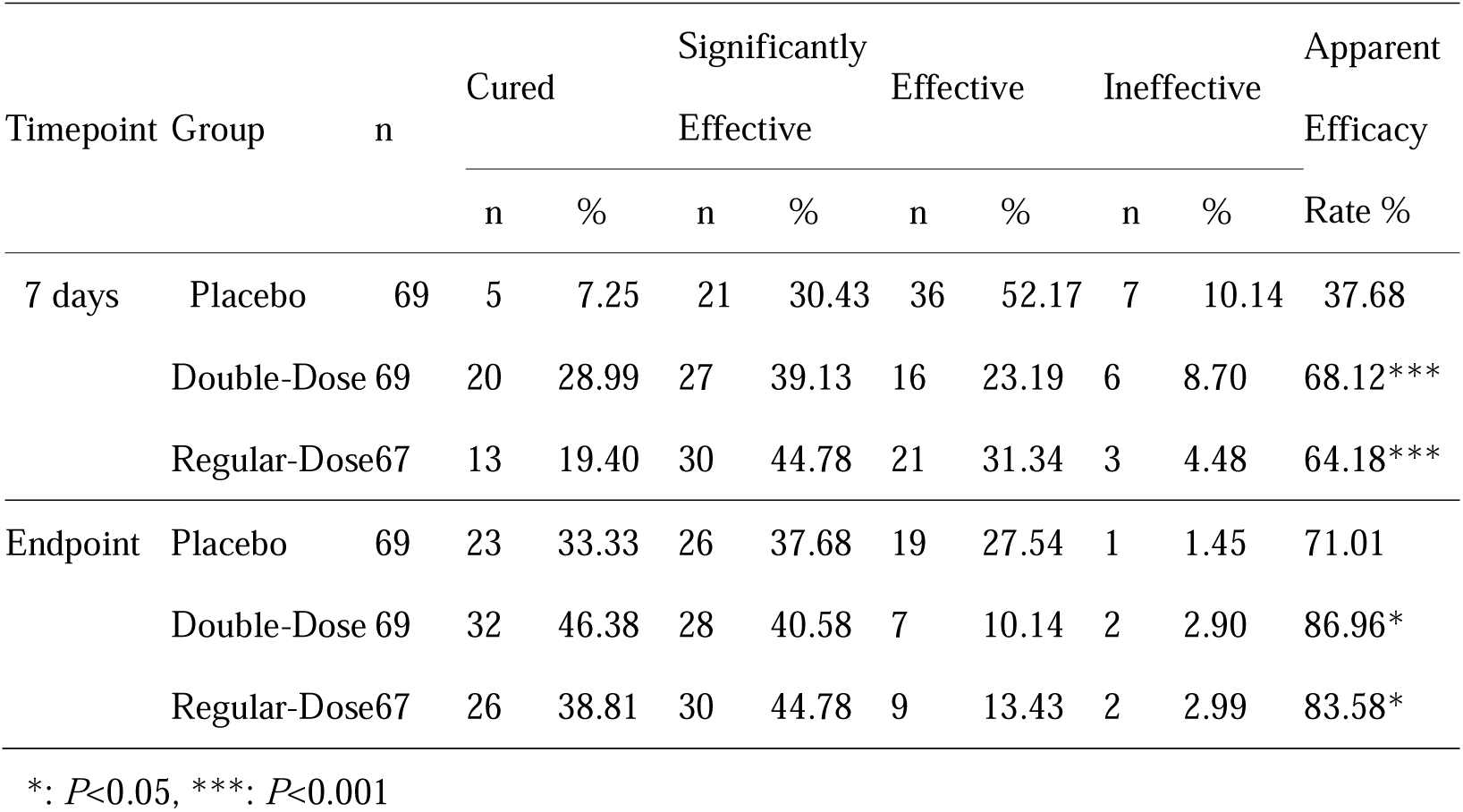
Comparison of TCM syndrome in three groups.

#### 3.1.6 Primary clinical outcomes

Study endpoints: The median clinical recovery time was 5 days for the double-dose group, 6 days for the regular-dose group, and 8 days for the placebo group, with significant statistical differences between groups in the order double-dose < regular-dose < placebo group. Both FAS and PPS analyses concluded consistently. The impact of the duration of illness at enrollment on clinical recovery times was assessed using COX regression analysis, showing statistically significant differences between groups, with consistent conclusions in both FAS and PPS analyses.

Three days into treatment, the clinical cure rates analyzed in the FAS were 7.25% for the placebo group, 5.80% for the double-dose group, and 10.45% for the regular-dose group; the differences were not statistically significant, with consistent conclusions in FAS and PPS analyses. After seven days of treatment, the clinical cure rates analyzed in the FAS were 46.38% for the placebo group, 75.36% for the double-dose group, and 73.13% for the regular-dose group, with significant statistical differences in the order double-dose group > regular-dose group > placebo group. Both FAS and PPS analyses concluded consistently. At the treatment endpoint (8-14 days), the clinical cure rates analyzed in the FAS were 97.10% for both the placebo and double-dose groups, and 95.52% for the regular-dose group; the differences were not statistically significant, with consistent conclusions in both FAS and PPS analyses.

The FAS analysis of the median time to complete defervescence for the three groups showed that the placebo group was at 5.0 (4.0, 6.0) days, the double-dose group at 3.0 (2.0, 4.0) days, and the regular-dose group at 3.0 (2.0, 4.0) days, with the order being double-dose < regular-dose < placebo group. There were statistically significant differences between groups, and the conclusions of the FAS and PPS analyses were consistent. The FAS analysis of the median time to relief from cough for the three groups found that the placebo group was at 7.0 (5.0, 9.0) days, the double-dose group at 5.0 (3.0, 5.0) days, and the regular-dose group at 5.0 (3.0, 7.0) days, with both the double-dose and regular-dose groups < placebo group. The differences between groups were statistically significant, and the conclusions of the FAS and PPS analyses were consistent. The FAS analysis of the median time to sputum relief for the three groups showed that the placebo group was at 7.0 (5.0, 8.0) days, the double-dose group at 4.0 (2.0, 5.0) days, and the regular-dose group at 5.0 (3.0, 6.0) days, with the order being double-dose group < regular-dose group < placebo group. There were statistically significant differences between groups, and the conclusions of the FAS and PPS analyses were consistent.

#### 3.1.7 Comparison of chest X-ray improvement

In this study, the regular-dose group reported one case that progressed to severe CAP or refractory MPP. The incidence of progression to severe CAP or refractory MPP showed no statistically significant differences between groups, and the conclusions of the FAS and PPS analyses were consistent. The FAS analysis of chest X-ray improvements at the treatment endpoint revealed that the placebo group had a normalization rate of 26.56%, a significant improvement rate of 40.63%, and an improvement rate of 14.06%, totaling an improvement rate of 81.25%. The double-dose group had a normalization rate of 24.24%, a significant improvement rate of 53.03%, and an improvement rate of 18.18%, totaling an improvement rate of 95.45%. The regular-dose group had a normalization rate of 24.19%, a significant improvement rate of 51.61%, and an improvement rate of 19.35%, totaling an improvement rate of 95.16%. The overall chest X-ray improvement rates were in the order: double-dose group > regular-dose group > placebo group, with statistically significant differences observed, and the conclusions of the FAS and PPS analyses were consistent.

#### 3.1.8 Effectiveness in TCM syndrome

The FAS analysis of the apparent rate of improvement in TCM syndrome at 7 days of treatment showed that the placebo group was at 37.68%, the double-dose group at 68.12%, and the regular-dose group at 64.18%, with the order being double-dose > regular-dose > placebo group. The differences were statistically significant, and the conclusions of the FAS and PPS analyses were consistent. At the treatment endpoint, the apparent rate of improvement in TCM syndrome for the placebo group was 71.01% (72.73%), for the double-dose group 86.96% (89.39%), and for the regular-dose group 83.58% (85.71%), with the order being double-dose > regular-dose > placebo group. The differences were statistically significant, and the conclusions of the FAS and PPS analyses were consistent.

#### 3.1.9 Secondary clinical outcomes

Study participants included those diagnosed with Phlegm-Heat Obstructing Lung Syndrome and Toxic-Heat Obstructing Lung Syndrome. The proportions of participants with Phlegm-Heat Obstructing Lung Syndrome in each group were 94.20%, 95.65%, and 92.54%, respectively. Subgroup analysis for participants with Phlegm-Heat Obstructing Lung Syndrome included:

Median Clinical Cure Time in FAS Analysis: The placebo group was at 8.0 (6.0, 9.0) days, the double-dose group at 5.0 (5.0, 7.0) days, and the regular-dose group at 6.0 (5.0, 8.0) days, with the order being double-dose < regular-dose < placebo group. Differences between groups were statistically significant, with consistent conclusions in both FAS and PPS analyses.

Overall Chest X-Ray Improvement Rate at Treatment Endpoint in FAS Analysis: The placebo group was at 80.00%, the double-dose group at 95.24%, and the regular-dose group at 94.74%, with the order being double-dose > regular-dose > placebo group. Differences were statistically significant, with consistent conclusions in both FAS and PPS analyses.

TCM Syndrome Apparent Efficacy Rate at 7 Days: The placebo group was at 36.92%, the double-dose group at 66.67%, and the regular-dose group at 62.90%, with the order being double-dose > regularose > placebo group. Differences were statistically significant, with consistent conclusions in both FAS and PPS analyses.

TCM Syndrome Apparent Efficacy Rate at Treatment Endpoint: The placebo group was at 69.23%, the double-ose group at 86.36%, and the regular-dose group at 82.26%, with the order being double-dose > regular-dose > placebo group. Differences were statistically significant, with consistent conclusions in both FAS and PPS analyses.

#### 3.1.10 Safety

The Safety Set included 205 patients (69 in the placebo group, 69 in the double-dose group, and 67 in the regular-dose group). Researchers reported 41 adverse events (AEs) occurring 48 times, with the placebo group reporting 15 cases (21.74%) occurring 19 times, the double-dose group reporting 6 cases (8.70%) occurring 8 times, and the regular-dose group reporting 20 cases (29.85%) occurring 21 times. The incidence of AEs showed statistically significant differences between groups. In this clinical study, a total of 2 serious adverse events (SAEs) were reported, occurring 2 times: the placebo group reported 0 cases (0.00%) occurring 0 times; the double-dose group reported 1 case (1.45%) occurring 1 time, due to bacterial infection; the regular-dose group reported 1 case (1.49%) occurring 1 time, due to right-sided pleural effusion. The incidence of SAEs showed no statistically significant differences between groups.

Within all reported adverse events, researchers identified 8 adverse drug reactions (ADRs) occurring 10 times. In the placebo group, 5 cases (7.25%) occurred 7 times, including 2 cases of elevated alanine aminotransferase, 1 case of elevated aspartate aminotransferase, 1 case of an abnormal electrocardiogram, 1 case of abdominal pain, and 2 cases of diarrhea. The double-dose group reported 0 cases (0.00%) occurring 0 times. The regular-dose group reported 3 cases (4.48%) occurring 3 times, including 1 case of pleural effusion, 1 case of allergic dermatitis, and 1 case of agranulocytosis. The incidence of ADRs showed no statistically significant differences between groups.

## 4. Discussion

MPP is a prevalent type of pneumonia among school-aged children, constituting 10-40% of CAP cases in this age group. Effective treatment aims to reduce severe cases, prevent deaths, and minimize long-term sequelae. TCM, specifically JZOL, has demonstrated clinical efficacy in various stages of MPP by inhibiting MP activity, modulating inflammatory responses, and enhancing immune function. Based on network target [51, 52], this study aims to uncover the mechanism of the combination of JZOL and AZ against MPP, and further evaluate the synergistic effects of combining JZOL with AZ to shorten disease duration, alleviate symptoms, improve prognosis, and determine the optimal clinical dosage and safety in treating MPP in children.

The study revealed that the combined treatment of JZOL and AZ works through a modular mechanism by targeting several biological pathways based on network target [53]. These include hormone response, cell differentiation and migration, signal transduction, and oxygen and hypoxia response. Key signaling pathways involved are TNF, PI3K-AKT, and MAPK, which play crucial roles in inflammation, immune response, and pathogen clearance in MPP. The hormone response module highlights the role of corticosteroids in rapidly improving clinical symptoms. Cell differentiation and migration involve EMT crucial for tissue remodeling. The oxygen response module addresses hypoxia-related complications like atelectasis and pleural effusion. The clinical trial results showed that the double-dose JZOL group had the shortest median clinical recovery time (5 days) compared to the regular-dose group (6 days) and placebo group (8 days). Additionally, the double-dose group exhibited the highest clinical cure rates and significant chest X-ray improvements. The combination therapy significantly improved TCM syndrome scores and demonstrated a safe profile with no significant differences in serious adverse events among the groups.

In the treatment of MPP, western medicine mainly focuses on anti-MP, with macrolide antibiotics being the first choice, among which AZ is the preferred treatment, while TCM has the characteristics of “multiple components and multiple targets”. Our research has found that the combination of JZOL and AZ does not act on a single target against MPP, but on network target. MAPK, JNK, and other cytokines can bind to TLR2 and be activated during Mycoplasma pneumoniae infection, inducing nuclear translocation of NF-κB and phosphorylation of PI3K/Akt in airway epithelial cells, leading to a significant increase in IL-8 production. In addition, the increase of TNF-α in the submucosal layer during Mycoplasma pneumoniae infection interacts with TNF-R on mast cells, leading to the release of a large amount of mediators, increased mucus production, and airway hyperresponsiveness. These network targets play a key role in the pathophysiology of MPP.

There were also some limitations in this study. First, based on network target analysis, we uncovered the mechanism of the combination of JZOL and AZ against MPP, in which the modular regulatory effects center-mediated by TNF, MAPK, and PI3K-Akt signaling pathways played imported roles. However, verifications in vivo and in vitro were also needed in further studies. And we were not able to collected clinical samples from children patients. Fortunately, we have validated our molecular level findings on public transcriptomics datasets at the compounds’ level owing to the more abundant resources of transcriptomics databases.

In conclusion, our study demonstrated that the combination of JZOL and AZ provides a significant synergistic effect in treating MPP in children. The network pharmacology analysis indicated that this combination targets multiple signaling pathways such as TNF, PI3K-AKT, and MAPK, which are crucial for inflammation, immune response, and pathogen clearance. Clinically, the combined therapy significantly shortened recovery time and improved cure rates, with a favorable safety profile. This integrative approach highlights the benefits of combining TCM with Western medicine, enhancing overall efficacy and safety, and offering new insights into the management of pediatric MPP through the synergy of both medical systems.

## Data Availability

All data produced in the present study are available upon reasonable request to the authors.

## Author contributions

Chengliang Zhong and Shengxuan Guo provided data interpretation and drafted the manuscript. Qingyuan Liu and Boyang Wang performed the computational analysis and drafted the manuscript. Boyang Wang and Deyang Sun performed the biological analysis and drafted the manuscript. Siyuan Hu conducted scheme design and provided technical support. Xinmin Li, Ying Ding, Bin Yuan, Jing Liu, Long Xiang, Nan Li, Zheng Xue, Yan Li, Yiqun Teng, and Rongsong Yi screened the enrolled patients and collected clinical data. Shao Li supervised and guided the study. Rong Ma conducted scheme design, data interpretation, and manuscript revision.

## Data sharing statement

Data available on request from authors

## Acknowledgements

This work was supported by the Jiangsu Provincial Department of Science and Technology-Basic Research Program Natural Science Fund - Frontier Leading Technology Basic Research Special Project “Research on the efficacy substance of traditional Chinese medicine compound based on the combination of disease and syndrome” (SBK2023050003).

## Abbreviations

ADR: adverse drug reactions
AE: adverse event
AZ: azithromycin
CI: confidence interval
FAS: full analysis set
JZOL: Jinzhen Oral Liquid
MP: Mycoplasma pneumoniae
MPP: Mycoplasma pneumoniae pneumonia
PPS: per protocol set
RMPP: refractory M. pneumoniae pneumonia
SAE: serious adverse event
SMPP: severe Mycoplasma pneumoniae pneumonia
SS: safety set
TCM: traditional Chinese medicine

## Notes

### Competing Interest Statement

The authors have declared no competing interest.

### Clinical Trial

The research has been registered with the China Clinical Trials Registry (registration number: ChiCTR1800019007) and has passed the ethical review of the leading unit's medical ethics committee (ethics approval number: TYLL2017[Y]020).

### Author Declarations

The research has been registered with the China Clinical Trials Registry (registration number: ChiCTR1800019007) and has been approved by the Medical Ethics Committee of the First Affiliated Hospital of Tianjin University of Traditional Chinese Medicine, which is the ethical review of the leading unit's medical ethics committee (ethics approval number: TYLL2017[Y]020).

